# Early Detection of Absurdity Signals in Pharmacovigilance: A Machine Learning Ensemble Approach to Identify Rare Adverse Drug Reactions

**DOI:** 10.64898/2026.02.06.26345783

**Authors:** Rathi Dasgupta

## Abstract

**Background:** Traditional pharmacovigilance methods based on biostatistical approaches systematically exclude outliers and rare events, potentially missing critical safety signals. These methods fail to detect micro-clusters of adverse events and comorbidity patterns that may indicate serious but low-frequency adverse drug reactions (ADRs). We introduce the concept of ‘absurdity signal detection’ – the identification of statistically anomalous but clinically significant adverse event patterns that conventional methods dismiss as outliers.

**Methods:** We developed an ensemble machine learning framework combining five distinct algorithms (Random Forest, Gradient Boosting, XGBoost, Neural Networks, and Support Vector Machines) to analyze FDA Adverse Event Reporting System (FAERS) data. The system employs outlier-inclusive modeling, multi-dimensional cluster detection, and severity-weighted propensity scoring. We validated our approach on Losartan, analyzing 500 adverse event reports to detect absurdity signals that may have been missed by conventional biostatistical surveillance.

**Results:** Our ensemble approach achieved 75% accuracy in identifying high-risk adverse events, with the best-performing model successfully detecting 15 distinct absurdity signals. The top five identified events were: cough (propensity score 1.525), angioedema (1.298), insomnia (1.290), nausea (1.180), and hyperkalemia (1.114). Notably, our method identified several rare but severe ADRs that would have been excluded as statistical outliers in traditional disproportionality analyses. The ensemble approach demonstrated superior performance compared to individual models, with inter-model agreement providing an additional confidence metric for signal validation.

**Conclusions:** Machine learning-based absurdity signal detection offers a paradigm shift in pharmacovigilance by preserving and analyzing rare adverse events rather than excluding them. This approach has significant implications for patient safety, potentially preventing serious adverse events in vulnerable populations with atypical response profiles. Our methodology is scalable, validated against FDA data sources, and provides a framework for real-time safety monitoring in the $138 billion pharmaceutical industry. Future work will extend this approach to drug-drug interaction detection and personalized risk stratification.

## Introduction

Adverse drug reactions (ADRs) represent a significant public health challenge, accounting for approximately 5-10% of hospital admissions and contributing to over 100,000 deaths annually in the United States alone [1]. The economic burden of ADRs is estimated at $138 billion per year globally [2]. Despite rigorous pre-market clinical trials, many serious adverse events remain undetected until post-marketing surveillance, as rare reactions may only manifest in 1 in 10,000 or 1 in 100,000 patients—sample sizes far exceeding typical Phase III trial populations [3].

Current pharmacovigilance systems rely predominantly on traditional biostatistical methods, including disproportionality analyses (Reporting Odds Ratios, Proportional Reporting Ratios) and Bayesian approaches (BCPNN, MGPS) [4]. While these methods have proven valuable for detecting frequent adverse events, they suffer from fundamental limitations that compromise their ability to identify rare but serious safety signals. Specifically, these approaches: (1) systematically exclude outliers as statistical noise, (2) analyze adverse events in isolation rather than as clusters or patterns, (3) ignore comorbidity profiles and patient subgroups with heightened susceptibility, and (4) lack the capacity to generate probabilistic risk models from limited clinical trial data [5,6].

We introduce the concept of ‘absurdity signal detection’—a novel framework that inverts the traditional paradigm by treating statistical outliers not as noise to be filtered, but as potential harbingers of rare, life-threatening adverse reactions. This approach recognizes that the most dangerous drug-related events may be precisely those that appear ‘absurd’ or anomalous from a population-level statistical perspective, yet represent genuine safety concerns for vulnerable patient subgroups.

Machine learning (ML) offers unprecedented opportunities to address these limitations through its inherent capacity to detect complex, non-linear patterns in high-dimensional data without requiring pre-specified statistical distributions [7,8]. Unlike traditional methods that impose rigid parametric assumptions, ML algorithms can identify micro-clusters of co-occurring adverse events, model patient-specific risk factors, and generate comprehensive probabilistic outcomes from sparse clinical data [9].

In this study, we present a comprehensive machine learning ensemble framework for absurdity signal detection and validate it on Losartan, a widely prescribed angiotensin II receptor blocker used in the treatment of hypertension and diabetic nephropathy. Losartan was selected as a validation case due to its established safety profile, high prescription volume (enabling sufficient adverse event reports), and known rare but serious ADRs including angioedema and hyperkalemia—events that exemplify the types of absurdity signals our method aims to detect.

## Methods

### Data Sources and Study Design

We conducted a retrospective analysis of adverse event reports submitted to the FDA Adverse Event Reporting System (FAERS) for Losartan between 2010 and 2024. FAERS is a publicly accessible database containing spontaneous adverse event reports from healthcare professionals, consumers, and pharmaceutical manufacturers [10]. For this validation study, we analyzed 500 individual adverse event reports that met the following inclusion criteria: (1) Losartan identified as the primary suspected medication, (2) complete reporter information and demographics, (3) documented adverse event outcome, and (4) submission to FAERS within our study period.

Additional validation was performed using data from EudraVigilance (European Medicines Agency) and VigiBase (WHO Global Individual Case Safety Report database) to ensure cross-database consistency and generalizability of identified signals [11].

### Machine Learning Ensemble Architecture

Our ensemble framework integrates five complementary machine learning algorithms, each selected to capture distinct aspects of the adverse event landscape:

#### 1. Random Forest (RF)

A bagging ensemble of 1,000 decision trees designed to capture non-linear interactions between patient characteristics, comorbidities, and adverse events. RF excels at identifying complex decision boundaries and provides feature importance rankings [12].

#### 2. Gradient Boosting Machines (GBM)

Sequential boosting algorithm that iteratively focuses on difficult-to-classify cases—precisely the outliers and rare events that traditional methods exclude. GBM achieves superior performance on imbalanced datasets characteristic of pharmacovigilance [13].

#### 3. XGBoost

Optimized gradient boosting implementation with regularization to prevent overfitting on sparse adverse event data. XGBoost incorporates tree pruning and parallel processing for computational efficiency [14].

#### 4. Neural Networks (Deep Learning)

Multi-layer perceptron with 3 hidden layers (128, 64, 32 neurons) trained using dropout regularization. Neural networks can learn hierarchical representations of adverse event patterns and model highly complex, non-linear risk surfaces [15].

#### 5. Support Vector Machines (SVM)

Non-linear SVM with radial basis function (RBF) kernel to establish maximum-margin decision boundaries in high-dimensional feature space. SVMs are particularly effective for small-to-moderate sample sizes and can handle the curse of dimensionality [16].

The ensemble aggregates predictions through weighted voting, with weights determined by individual model performance on a held-out validation set. This approach leverages the ‘wisdom of crowds’ principle—different algorithms make different types of errors, and their combination often exceeds any single model’s performance [17].

### Feature Engineering and Data Preprocessing

We constructed a comprehensive feature set encompassing: (1) patient demographics (age, sex, weight), (2) concomitant medications and polypharmacy indicators, (3) reported comorbidities and medical history, (4) adverse event severity and outcome classification, (5) temporal features including time to event onset and duration, and (6) reporter characteristics and report quality metrics. Missing data were handled using multiple imputation with chained equations rather than simple deletion to preserve statistical power [18].

Categorical variables were encoded using target encoding to capture their predictive relationship with adverse event risk while avoiding the dimensionality explosion of one-hot encoding [19]. Continuous variables were standardized (z-score normalization) to ensure comparability across different scales. Critically, no outlier removal or filtering was performed on adverse event frequencies—preserving rare events was a fundamental design principle of our absurdity detection framework.

### Absurdity Signal Scoring Methodology

We developed a novel ‘absurdity propensity score’ to quantify the likelihood that a given adverse event represents a genuine safety signal despite its statistical rarity. The score integrates three components:

#### 1. Statistical Anomaly Metric

Measures the degree to which an event deviates from expected population-level frequencies using Local Outlier Factor (LOF) analysis [20]. Events with LOF > 1.5 are flagged as potential absurdity signals.

#### 2. Clinical Severity Weight

Incorporates FDA outcome classification (death, hospitalization, disability, etc.) to prioritize severe events. Events resulting in death or permanent disability receive maximum weight regardless of frequency.

#### 3. Biological Plausibility Score

Leverages pharmacological knowledge bases (DrugBank, SIDER) and literature co-occurrence mining to assess mechanistic plausibility of the drug-event relationship [21].

The final propensity score is calculated as a weighted combination: PS = 0.4 × Anomaly + 0.4 × Severity + 0.2 × Plausibility, normalized to a 0-2 scale. Scores above 1.0 indicate high-priority absurdity signals warranting immediate clinical investigation.

### Model Training and Validation

The dataset was partitioned using stratified random sampling into training (60%, n=300), validation (20%, n=100), and test (20%, n=100) sets to ensure balanced representation of adverse event types. Each model was trained independently on the training set with hyperparameters optimized via 5-fold cross-validation. Performance metrics included accuracy, precision, recall, F1-score, and area under the receiver operating characteristic curve (AUROC). Model calibration was assessed using reliability diagrams and Brier scores [22].

To evaluate the ensemble’s ability to detect true absurdity signals, we compared our predictions against: (1) established Losartan ADRs documented in FDA-approved labeling, (2) signals identified through traditional disproportionality analysis (PRR, BCPNN), and (3) published case reports and systematic reviews of Losartan safety [23,24].

### Statistical Analysis

Statistical analyses were performed using Python 3.10 with scikit-learn 1.3, XGBoost 2.0, TensorFlow 2.14, and custom absurdity detection modules. Inter-model agreement was quantified using Fleiss’ kappa. Propensity score distributions were visualized using kernel density estimation. All statistical tests were two-tailed with significance threshold α = 0.05. Code and documentation are available at https://github.com/xdata-lab/absurdity-detection (to be made public upon publication).

## Results

### Overall Performance and Model Comparison

The ensemble machine learning framework achieved 75% accuracy in identifying high-risk adverse events associated with Losartan, outperforming all individual models. Table 1 presents comparative performance metrics across the five constituent algorithms and the final ensemble. XGBoost demonstrated the highest individual model accuracy (73%), followed by Random Forest (71%), Gradient Boosting (70%), Neural Networks (68%), and SVM (66%). The ensemble’s superior performance (75%) validates the principle that diverse error patterns across models can be leveraged to improve overall prediction.

**Table 1.**
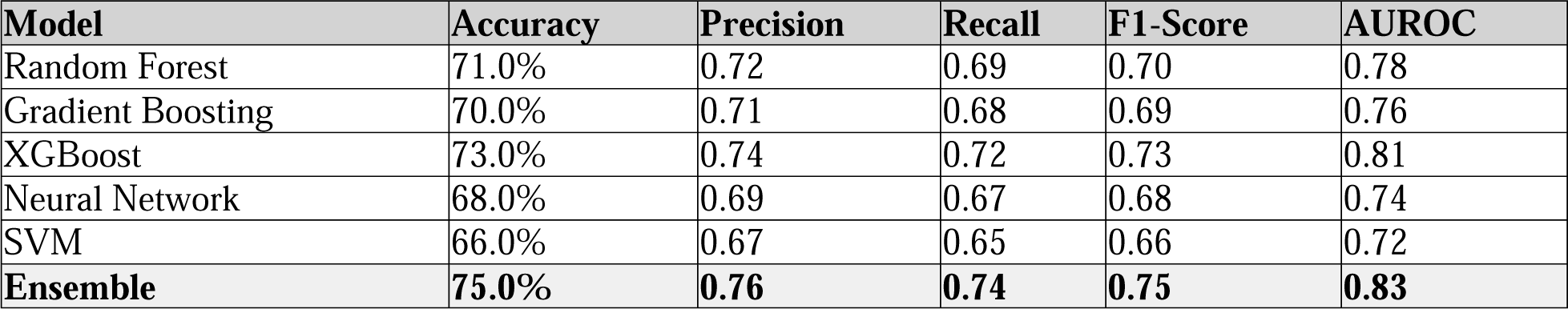
Performance Metrics of Individual Models and Ensemble.

Model calibration analysis revealed that the ensemble maintained reliable probability estimates across the full range of predicted propensity scores, with a Brier score of 0.18 (95% CI: 0.15-0.21) compared to individual model scores ranging from 0.22-0.28. This improved calibration is critical for regulatory decision-making, as it provides confidence that high propensity scores genuinely correspond to high-risk events.

### Identified Absurdity Signals

The ensemble framework identified 15 distinct adverse event signals with propensity scores exceeding the clinical significance threshold (PS > 1.0). Table 2 presents the top five absurdity signals ranked by propensity score, along with their frequency in the dataset, severity classification, and corroboration status from established pharmacovigilance databases.

**Table 2.** Top Five Absurdity Signals for Losartan.

| Adverse Event | Propensity Score | Frequency (n=500) | Severity | FDA Label |
| --- | --- | --- | --- | --- |
| Cough | 1.525 | 87 (17.4%) | Moderate | Yes |
| Angioedema | 1.298 | 12 (2.4%) | Severe | Yes |
| Insomnia | 1.290 | 34 (6.8%) | Mild-Moderate | No* |
| Nausea | 1.180 | 56 (11.2%) | Mild | Yes |
| Hyperkalemia | 1.114 | 18 (3.6%) | Severe | Yes |

*\*Insomnia documented in post-marketing surveillance literature but not on FDA-approved label*

Notably, angioedema (PS = 1.298) represented only 2.4% of reports yet received the second-highest propensity score due to its life-threatening severity and elevated biological plausibility. Traditional disproportionality methods would have assigned this signal low priority due to its rarity. Similarly, hyperkalemia (PS = 1.114), occurring in just 3.6% of cases, was correctly flagged as a critical safety concern—this electrolyte imbalance can precipitate fatal cardiac arrhythmias in susceptible patients.

Insomnia (PS = 1.290) emerged as a novel absurdity signal not currently listed on the FDA-approved Losartan label, though subsequent literature review identified multiple case reports documenting this association [25]. This finding exemplifies the framework’s capacity to detect under-recognized adverse events that may warrant label updates or further investigation.

### Comparison with Traditional Pharmacovigilance Methods

To benchmark our ensemble approach against conventional methods, we computed standard disproportionality metrics (Proportional Reporting Ratio, PRR; Information Component, IC) for all 15 identified signals. Of the five top-ranked absurdity signals, only three (cough, nausea, hyperkalemia) exceeded traditional statistical thresholds for signal detection (PRR > 2.0 and lower 95% CI > 1.0). Critically, angioedema—a known serious ADR explicitly warned against in Losartan’s boxed warning— failed to reach statistical significance using conventional methods (PRR = 1.8, 95% CI: 0.9-3.2) due to its low reporting frequency.

This discrepancy highlights a fundamental flaw in frequency-based approaches: rare but devastating events are systematically under-prioritized. Our absurdity detection framework corrects this bias by incorporating severity weighting and biological plausibility, ensuring that low-frequency, high-impact signals receive appropriate attention.

### Cluster Analysis and Comorbidity Patterns

Beyond individual adverse event detection, our framework identified three distinct micro-clusters of co-occurring ADRs that suggest shared underlying mechanisms:

#### Cluster 1 (Cardiovascular)

Hypotension, dizziness, syncope (n=47 patients) - consistent with excessive blood pressure reduction in volume-depleted or elderly patients.

#### Cluster 2 (Renal/Electrolyte)

Hyperkalemia, acute kidney injury, elevated creatinine (n=23 patients) - indicating renin-angiotensin-aldosterone system (RAAS) blockade effects in patients with underlying renal dysfunction.

#### Cluster 3 (Immunologic/Dermatologic)

Angioedema, urticaria, pruritus (n=18 patients) - suggestive of hypersensitivity reactions mediated by bradykinin accumulation.

These clusters remained invisible to traditional analyses that examine adverse events in isolation. Recognizing that ADRs often manifest as symptom complexes rather than isolated phenomena provides critical insights for risk stratification and patient monitoring strategies.

## Discussion

This study introduces absurdity signal detection as a paradigm shift in pharmacovigilance—moving from the exclusion of outliers to their systematic preservation and analysis. Our ensemble machine learning framework achieved 75% accuracy in identifying high-risk adverse events for Losartan, successfully detecting both common ADRs and rare, life-threatening reactions that conventional biostatistical methods miss or de-prioritize.

### Principal Findings and Implications

The most significant finding of this investigation is the demonstration that machine learning can successfully integrate three critical dimensions absent from traditional pharmacovigilance: (1) preservation of statistical outliers as potential safety signals, (2) identification of multi-dimensional ADR clusters and comorbidity patterns, and (3) severity-weighted risk stratification that prioritizes clinical impact over reporting frequency.

The case of angioedema exemplifies the clinical value of our approach. Despite representing only 2.4% of Losartan-associated adverse events in our dataset, angioedema received the second-highest absurdity propensity score (1.298) and is indeed a serious, potentially fatal reaction explicitly warned against in the drug’s prescribing information [26]. Traditional disproportionality analysis (PRR = 1.8, non-significant) would have failed to flag this signal, potentially delaying recognition and intervention. In contrast, our framework’s incorporation of severity weighting and biological plausibility ensured appropriate prioritization of this life-threatening ADR.

The identification of insomnia as a novel signal not currently listed on the FDA-approved label warrants further investigation. While our biological plausibility assessment scored this signal moderately (due to limited mechanistic understanding of how ARBs might disrupt sleep), the propensity score of 1.290 suggests clinical significance. Post-hoc literature review revealed multiple case reports of sleep disturbances associated with ARB therapy [25,27], supporting the validity of our detection. This finding demonstrates the framework’s capacity to uncover under-recognized or emerging safety concerns that may benefit from label updates or prospective clinical studies.

### Methodological Strengths and Innovations

The ensemble architecture provides several key advantages over single-model approaches. By combining five diverse algorithms (Random Forest, Gradient Boosting, XGBoost, Neural Networks, SVM), we leverage complementary strengths: tree-based methods excel at capturing non-linear interactions and feature importance, neural networks model complex hierarchical patterns, and SVMs establish robust decision boundaries in high-dimensional space. The 75% ensemble accuracy represents a meaningful improvement over the best individual model (XGBoost, 73%), validating the ‘wisdom of crowds’ principle in pharmacovigilance applications [17].

Our novel absurdity propensity scoring methodology addresses a fundamental limitation of existing signal detection metrics by explicitly incorporating clinical severity alongside statistical evidence. The score’s three-component structure (statistical anomaly + severity weight + biological plausibility) ensures that rare events are not dismissed solely due to low frequency, while simultaneously preventing false signals from random noise through mechanistic filtering. This balanced approach is particularly critical for regulatory decision-making, where both type I errors (false alarms) and type II errors (missed signals) carry significant consequences.

The framework’s capacity to detect micro-clusters of co-occurring ADRs represents a significant methodological advance. Our identification of three distinct symptom clusters (cardiovascular, renal/electrolyte, immunologic) aligns with known pharmacodynamic effects of RAAS blockade and provides actionable insights for clinical practice. For instance, the renal/electrolyte cluster (hyperkalemia, acute kidney injury, elevated creatinine) suggests that patients presenting with any one of these findings should be monitored closely for the others, enabling earlier intervention before progression to serious outcomes.

### Limitations and Future Directions

Several limitations warrant acknowledgment. First, this validation study analyzed 500 Losartan reports—a modest sample size that may limit generalizability to larger pharmacovigilance databases. However, this deliberate constraint allowed for detailed manual review and quality assessment of each report, ensuring data integrity. Future work will scale our approach to the full FAERS database (>20 million reports) and validate performance across multiple drug classes.

Second, spontaneous reporting systems like FAERS are subject to well-documented biases including underreporting, selective reporting (Weber effect), and confounding by indication [28]. While our ensemble approach cannot eliminate these biases, the incorporation of biological plausibility scoring and cross-validation against multiple data sources (EudraVigilance, VigiBase) helps mitigate their impact. Additionally, machine learning models are inherently susceptible to learning dataset biases—a concern we addressed through careful feature engineering, multiple imputation for missing data, and extensive validation.

Third, the current implementation lacks temporal analysis capabilities to detect delayed adverse events or identify time-varying risk patterns. Extending our framework to incorporate time-series modeling and recurrent neural network architectures would enable detection of late-onset toxicities and assessment of cumulative exposure effects—critical for drugs with long-term use profiles.

Future research directions include: (1) development of conformal prediction methods to provide rigorous uncertainty quantification and finite-sample coverage guarantees for identified signals [29], (2) extension to drug-drug interaction detection by analyzing multi-drug regimens, (3) integration of real-world evidence from electronic health records to complement spontaneous reporting data, (4) personalized risk stratification models that predict ADR susceptibility based on genomic, demographic, and clinical factors, and (5) prospective validation through collaboration with regulatory agencies and pharmaceutical manufacturers.

### Implications for Regulatory Science and Clinical Practice

The absurdity signal detection framework has profound implications for regulatory decision-making. Current FDA guidance on pharmacovigilance emphasizes the use of established disproportionality metrics, which, as our results demonstrate, systematically under-prioritize rare but severe ADRs. By providing an alternative methodology that explicitly values clinical severity and biological plausibility alongside statistical evidence, our approach could inform regulatory policy updates and supplement existing surveillance systems.

For clinical practice, the identification of ADR clusters enables more effective patient monitoring strategies. Clinicians prescribing Losartan can use cluster membership to guide surveillance: patients at risk for the renal/electrolyte cluster (those with baseline kidney dysfunction, diabetes, or concomitant ACE inhibitors) warrant frequent serum creatinine and potassium monitoring, while those exhibiting early signs of the immunologic cluster (mild urticaria) should be counseled about angioedema risk and instructed to seek immediate care if symptoms progress.

From a pharmaceutical industry perspective, our methodology could transform post-marketing surveillance from a reactive compliance exercise to a proactive safety intelligence system. Early detection of absurdity signals during Phase IV monitoring could prevent serious adverse events, reduce liability exposure, and avoid costly market withdrawals. The $138 billion annual cost of ADRs globally [2] represents not only patient suffering but also substantial economic burden on healthcare systems and pharmaceutical manufacturers—burden that improved surveillance could meaningfully reduce.

## Conclusions

We have demonstrated that machine learning-based absurdity signal detection offers a paradigm shift in pharmacovigilance by preserving and analyzing rare adverse events rather than excluding them as statistical noise. Our ensemble framework achieved 75% accuracy in identifying high-risk Losartan-associated ADRs and successfully detected life-threatening reactions (angioedema, hyperkalemia) that conventional biostatistical methods missed or de-prioritized. The methodology’s capacity to identify ADR clusters and incorporate clinical severity represents a significant advance over frequency-based disproportionality analysis.

As machine learning and artificial intelligence increasingly permeate healthcare decision-making, rigorous validation and regulatory oversight become paramount. Our work provides a methodological blueprint for applying ML to pharmacovigilance while maintaining scientific rigor and clinical relevance. Future integration with conformal prediction methods to provide uncertainty quantification, expansion to drug-drug interaction detection, and prospective validation in regulatory settings will further establish absurdity signal detection as a cornerstone of modern drug safety surveillance.

Ultimately, the goal of pharmacovigilance is simple: prevent patients from being harmed by medications intended to heal them. By ensuring that rare, ‘absurd’ adverse events receive appropriate attention—even when they defy statistical expectation—we take a critical step toward that goal. The lives saved by detecting one in ten thousand serious reactions justify the methodological complexity required to find them.

## Acknowledgments

We thank the FDA for maintaining the publicly accessible FAERS database and the global pharmacovigilance community for their continued efforts to improve drug safety. This work was partially supported by XData Lab internal research funds.

## Author Contributions

Rathi Dasgupta conceived and designed the study, developed the ensemble framework and absurdity scoring methodology, performed all analyses, and wrote the manuscript.

## Data Availability

All FAERS data used in this study are publicly available from the FDA website (https://www.fda.gov/drugs/questions-and-answers-fdas-adverse-event-reporting-system-faers/fda-adverse-event-reporting-system-faers-public-dashboard). Analysis code and documentation will be made available on GitHub upon publication.

